# Use of Unofficial Newspaper Data for COVID-19 Death Surveillance

**DOI:** 10.1101/2020.09.09.20191569

**Authors:** Mazbahul G. Ahamad, Monir U. Ahmed, Byomkesh Talukder, Fahian Tanin

## Abstract

**Objective:** To highlight the critical importance of unofficially reported newspaper-based deaths from coronavirus disease 2019 (COVID-19)–like illness (CLI) together with officially confirmed death counts to support improvements in COVID-19 death surveillance.

**Methods:** Both hospital-based official COVID-19 and unofficial CLI death counts were collected from daily newspapers between March 8 and August 22, 2020. We performed both exploratory and time-series analyses to understand the influence of combining newspaper-based CLI death counts with confirmed hospital death counts on the trends and forecasting of COVID-19 death counts. An autoregressive integrated moving average–based approach was used to forecast the number of weekly death counts for six weeks ahead.

**Results:** Between March 8 and August 22, 2020, 2,156 CLI deaths were recorded based on newspaper reporting for a count that was 55% of the officially confirmed death count (n = 3,907). This shows that newspaper reports tend to cover a significant number of COVID-19 related deaths. Our forecast also indicates an approximate total of 406 CLI expected for the six weeks ahead, which could contribute to a total of 2,413 deaths including 2,007 confirmed deaths expected from August 23 to October 3, 2020.

**Conclusions:** Analyzing existing trends in and forecasting the expected number of newspaper-based CLI deaths indicates yet-unreported COVID-19 death counts, which could be a critical source to estimate provisional COVID-19 death counts and mortality surveillance.

**Public Health Implications:** Considering unofficial newspaper-based CLI death counts is essential to identify COVID-19 death severity and surveillance needs to advance public health research efforts to prepare appropriate response strategies for low- and middle-income countries.

## 1. INTRODUCTION

In many developed and developing countries, coronavirus disease 2019 (COVID-19) death counts, mortality surveillance, and reporting procedures have been growing concerns since the coronavirus began spreading globally in early 2020. These have considerable public health consequences, including poor pandemic response strategies based on inaccurate and low-quality data and forecasting. For pandemic mortality surveillance, reporting authorities have been advised to report “[any] death resulting from a clinically compatible illness in a probable or confirmed COVID-19 case,” according to the World Health Organization.^1^ Death-reporting authorities in developed countries, such as the Centers for Disease Control and Prevention of the United States, continue to track hospital data-based confirmed COVID-19deaths together with those attributed to COVID-19–like illness (CLI).^2^ However, many low- and middle-income countries (LMICs) such as Bangladesh lack appropriate death surveillance systems officially; thus, CLI and related influenza-like illness (ILI) death counts are not being surveyed and used to estimate a true COVID-19 mortality rate.^3,4^

Moreover, many LMICs have no competent mortality surveillance guidelines.^4,5^ For example, the reporting authority in Bangladesh only reports confirmed hospital deaths without officially considering any CLI or ILI adjusted CLI deaths, which is predominantly attributed to the absence of a comprehensive reporting guideline and surveillance system.^3^ The lack of CLI and adjustment with ILI death counts is a potential source biases regarding of death surveillance and mortality estimate, which undermines the validity of the death counts and mortality reporting. Developing a complete death surveillance system, based on official and unofficial sources, to guide reporting authorities is, therefore, critical to ensuring the existence of a true data-driven pandemic response and mitigation strategies.^4^

Newspaper report–based death surveillance is useful to identify CLI deaths where the prevalence of CLI is frequent, but these reports remain unidentified in hospital or official reports. In Bangladesh, CLI death counts are gathered and reported weekly based on daily newspaper information by the Centre for Genocide Studies (CGS).^6^ This unofficial approach of death reporting offers public health researchers and policy analysts the opportunity to use a supplementary source to understand CLI death counts along with possible total death counts.

**Based on above background, the** objective was to highlight the critical relevance of newspaper-based CLI death data to support improved death surveillance, especially in Bangladesh-like LMICs, by estimating and comparing both the actual and forecasted trends of official hospital and unofficial newspaper data.

## 2. METHODS

We obtained officially confirmed COVID-19 death count data from the Johns Hopkins Coronavirus Resource Center^7^ and unofficial newspaper-based CLI death counts data from the CGS.^6^ Confirmed death counts were established based on hospital data collected by the Institute of Epidemiology Disease Control and Research, whereas the CGS reviews newspapers daily to estimate CLI death counts. The review and death-count estimation involved the content analysis of relevant newspaper reports by retracing every case and removing duplications published in different daily newspapers (Ref).

We considered the sum of official and unofficial death counts as the “total” death count. An autoregressive integrated moving average (ARIMA) model prediction was conducted to predict the forecasted number of weekly^4,8^ confirmed, CLI, and total death counts for six weeks ahead (Appendix A). For further understanding, we can also assume that 30% of CLI deaths might be related to ILI because the exact percentage is not officially available and we can refer this as “ILI-adjusted CLI” deaths and the respective total as “ILI-adjusted total” deaths.

Because official statistics of coronavirus tests, total cases, and death counts do not present the complete picture of Bangladesh,^6^ the intention of the forecasting herein was to estimate and present simple trends only from a comparative analysis standpoint rather than accurately predicting respective death counts. Statistical analysis was carried out using Stata version 16.1.^9^

## 3. RESULTS

Between March 8 and August 22, 2020, a total of 3,907 confirmed COVID-19 deaths from hospital data together 2,156 officially unreported CLI deaths from newspapers (thus achieving a count that was 55% that of total confirmed deaths) were recorded. We observed an average of 156 (95% confidence interval: 105.4–207.2) confirmed deaths per week, and 86 (95% confidence interval: 59.5–113.0) CLI deaths per week (Table 1). A significant mean difference between the confirmed and CLI death counts (P < 0.01) was noted during the same period.

**Table 1:** Actual and forecasted mean confirmed and CLI death counts.

| COVID-19 death | Confirmed death<br>Mean (95% CI) | CLI death<br>Mean (95% CI) | *t* | *P*-value |
| --- | --- | --- | --- | --- |
| **Actual period:** Mar 8–Aug 22 | 156.3 (105.4–207.2) | 86.2 (59.5–113.0) | 2.514 | 0.008 |
| **Forecasted period:** Aug 23–Oct 03 | 334.5 (311.4–357.6) | 68 (52.5–83.5) | 24.668 | 0.000 |

*Notes:* CI = Confidence Interval; *t* = t-statistic; *P*-value = probability value.

Our forecast suggested a total of 2,007 confirmed deaths could be expected to occur in the six weeks ahead, which could increase to 2,413 when considering an additional 406 unreported CLI deaths expected from August 23 to October 3, 2020 (Fig. 1). This also showed a significant difference (P < 0.001) existed between the official and CLI death counts (Table 1). The difference might be significant when we consider ILI adjusted total death count (i.e., with a 30% reduction due to ILI deaths) instead of the total (sum of confirmed and CLI) death count.

**Figure 1:**
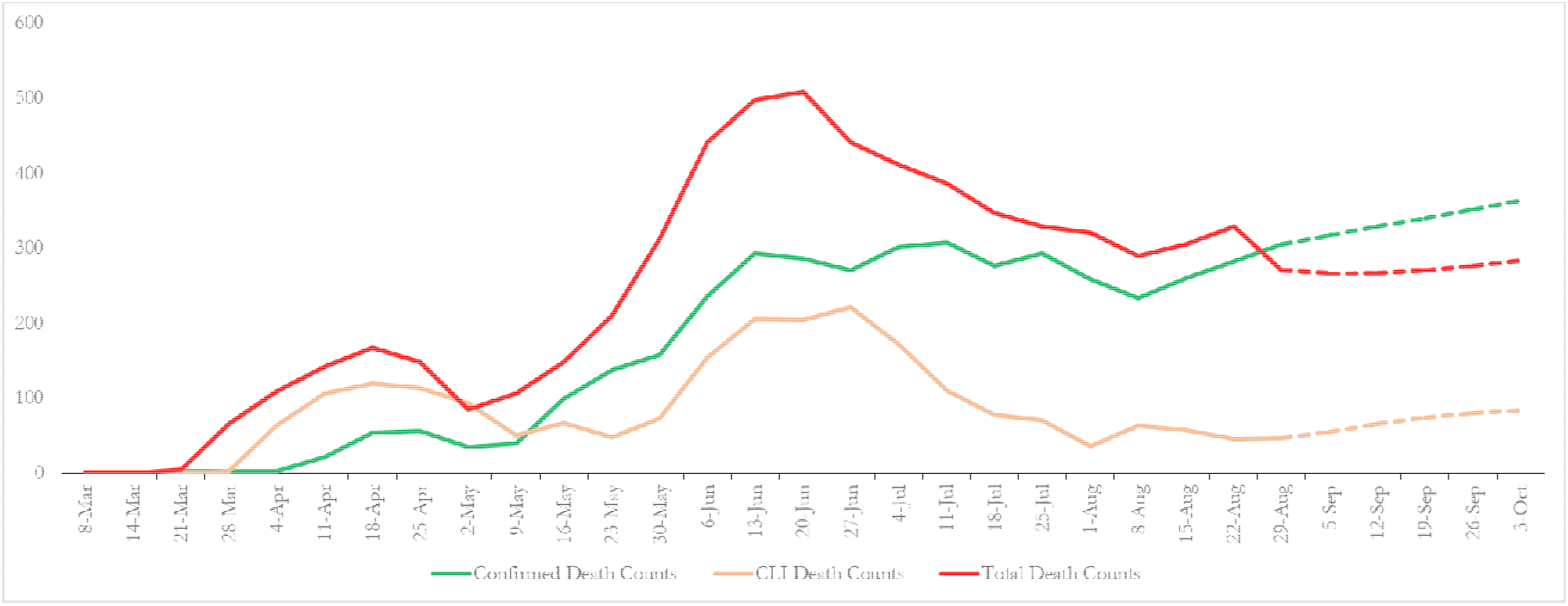
Actual and forecasted trends of confirmed, CLI, and total COVID-19 death counts in Bangladesh.

*Notes:* Solid lines present actual (Mar 8–Aug 22) death counts and dashed lines present forecasted (Aug 23–Oct 03) death counts.

## 4. DISCUSSION

During the COVID-19 pandemic, newspaper reports have covered a significant number of CLI deaths in Bangladesh. To the best of our knowledge, this paper includes the first empirical analysis to show how newspaper-based CLI data can produce important insights in countries wherein community-based death or mortality surveillance efforts are limited or nonexistent during the COVID-19 pandemic. Only officially confirmed hospital data are reported^7^ as part of mortality surveillance in Bangladesh, but newspaper-based CLI death reporting constitutes an alternative source of information^10^ in the absence of established community-based surveillance. This provided a more complete picture of the impact of COVID-19 on public health, particularly for CLI deaths, because many COVID-19 deaths occur outside of hospitals or other health facilities.

Newspaper-based CLI death counts offer a unique opportunity to strengthen death data surveillance and policy issues. First, the CLI death count reveals the excess death count^2,10^ related to CLI, which can be compared with pre-pandemic levels (e.g., by point-to-point or year-on-year basis). This is important for Bangladesh and other LMICs to be able to strengthen their existing COVID-19 mortality surveillance systems.

Second, we need the best possible forecasting to accurately predict the trajectory of the COVID-19 pandemic. Data availability and reliability are important concerns to maintain when predicting future waves. Using only officially confirmed death data might underestimate future death counts together with resource allocation priorities. So, newspaper data-based CLI death counts are a secondary source of public policy analysts in understanding clearer estimates when comparing with COVID-19–related death counts on a year-on-year basis. Given that most newspaper reports can be retrieved from electronic archives, interested researchers and policymakers in LMICs could estimate newspaper-based CLI death counts to facilitate and improve unofficial COVID-19 death surveillance.

### Limitations

The potential limitations of this research should be considered when adopting newspaper-based mortality surveillance. First, newspapers cannot cover all CLI death throughout a country if the number is very high as has been the case in Italy or the United States. Second, reported CLI deaths are not adjusted with ILI deaths and adjustment estimates and guidelines are unavailable in many LMICs, including Bangladesh. Third, very few LMICs have the institutional and other necessary capacities to rigorously analyze newspaper death-reporting, which may restrict them from adopting newspaper-based mortality assessment and surveillance.

### Public health implications

Because of significant reporting gaps between official and unofficial COVID-19 mortality data in many LMICs, policymakers and public health researchers should be concerned about confirmed, CLI, and ILI death counts. An inaccurate or incomplete understanding of these numbers could generate incorrect mortality and forecasting discrepancies and could affect pandemic preparedness and response strategies.

By developing an all-inclusive mortality data surveillance and reporting approach using both hospital and newspaper data, public health authorities could better inform and guide national response and recovery planning to limit the impact of COVID-19, which remains a major public health policy concern in many LMICs.

## CONTRIBUTORS

M. Ahamad conceptualized and designed the study. M. Ahamad and F. Tanin collected the data. M. Ahamad and M. Ahmed performed statistical analyses and data interpretation. M. Ahamad, M. Ahmed, B. Talukder, and F. Tanin reviewed and edited the manuscript and approved the final version.

## Data Availability

Data and codes used in the study will be available at https://dataverse.harvard.edu/dataverse/mazbahulahamad after publication.

## ACKNOWLEDGEMENTS

We thank the Bangladesh Peace Observatory (www.peaceobservatory-cgs.org/#/highlights), hosted at the CGS, University of Dhaka, in partnership with the United Nations Development Programme, for collecting and disseminating CLI death count data. We also acknowledge the official death statistics provided by the Institute of Epidemiology Disease Control and Research, Bangladesh (www.iedcr.gov.bd) and the Johns Hopkins Coronavirus Resource Center (www.coronavirus.jhu.edu). Data and codes used in the study will be available at https://dataverse.harvard.edu/dataverse/mazbahulahamad after publication.

## HUMAN PARTICIPATION PROTECTION

Ethical approval was not sought as this study used publicly available data.

## Supplementary Material

### S1: Forecasting of officially confirmed, newspaper-based CLI, and total COVID-19 death counts

We used an ARIMA technique to forecast the officially confirmed, newspaper-based CLI, and total death counts (sum of confirmed and CLI) for six weeks ahead. An ARIMA model typically uses an iterative process of identifying a practically useful model from a standard category of models without presuming any pattern in the historical data of the variable. Both autocorrelation and partial autocorrelation functions were used for the identification of the stationary in an ARIMA model. When the correlogram does not diminish in the face of large lags, it is assumed that nonstationarity is present. Additionally, partial autocorrelation is typically used to test for the presence of any unit root in the series.

An ARIMA model considers either an autoregressive (AR) or moving average (MA) or both. If the partial autocorrelation function (PACF) of the differenced variable presents a sharp cutoff and/or the lag of one autocorrelation indicates positive, then we can add one or more AR terms to the specific model. The number of AR terms explains the lag beyond the cutoff period of PACF. If the autocorrelation function (ACF) of the differenced variable presents a sharp cutoff and/or the lag of one autocorrelation indicates negative, then we can add one or additional MA term to the specific model. The lag beyond which the ACF cuts off is the indicated number of MA terms. A detailed description can be found at https://people.duke.edu/~rnau/411arim3.htm. Based on these criteria, we forecasted each series using an automatic ARIMA forecasting technique (Table S1).

**Table SI:**
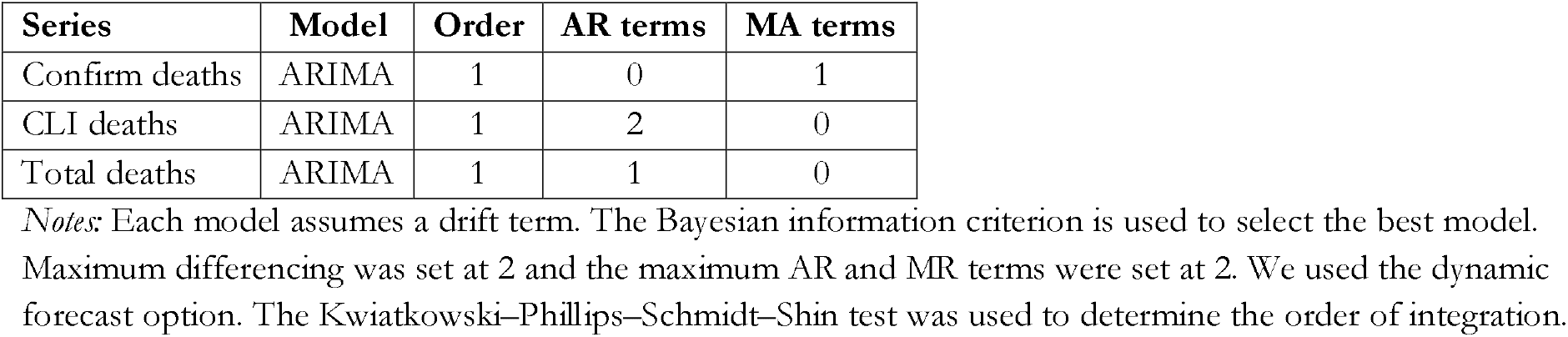
Description of ARIMA models.

## REFERENCES

1 World Health Organization (WHO). Coronavirus disease (COVID-19): Situation report - 102. Geneva, 2020 https://www.who.int/docs/default-source/coronaviruse/situation-reports/20200501-covid-19-sitrep.pdf?sfvrsn=742f4a18_4 (accessed July 19, 2020).

2 National Center for Health Statistics (NCHS). Technical notes: provisional death counts for coronavirus disease (COVID-19). 2020. https://www.cdc.gov/nchs/nvss/vsrr/covid19/tech_notes.htm.

3 Ahamad M, Tanin F, Talukder B. Confirmed and unreported COVID-19-like illness death counts: an assessment of reporting discrepancy. medRxiv 2020;2020.07.20.20158139.

4 Leon D, Shkolnikov V, Smeeth L, Magnus P, Pechholdová M, Jarvis C. COVID-19: a need for real-time monitoring of weekly excess deaths. Lancet. 2020; 395: e81.

5 Ihekweazu C, Agogo E. Africa’s response to COVID-19. BMC Med 2020; 18. DOI: 10.1186/s12916-020-01622-w.

6 Centre for Genocide Studies (CGS). Covid19graphics 17. Dhaka, 2020.

7 JHCRC. Johns Hopkins Coronavirus Resource Center. 2020. https://coronavirus.jhu.edu/ (accessed April 15, 2020).

8 Setel P, Abouzahr C, Atuheire EB, et al. Mortality surveillance during the COVID-19 pandemic. Bull. World Health Organ. 2020; 98: 374.

9 StataCorp. Stata Statistical Software: Release 16. College Station, TX: StataCorp LLC, 2019.

10 Woolf S, Chapman D, Sabo R, Weinberger D, Hill L. Excess deaths from COVID-19 and other causes, March-April 2020. JAMA 2020;E1–3.

